# Quantification of Conflicts of Interest in an Online Point-of-Care Clinical Support Website

**DOI:** 10.1101/19001859

**Authors:** Ambica C. Chopra, Stephanie Tilberry, Kaitlyn E. Sternat, Daniel Y. Chung, Stephanie D. Nichols, Brian J. Piper

## Abstract

Online medical reference websites are utilized by health care providers to enhance their education and decision making. However, these resources may not adequately reveal pharmaceutical-author interactions and their potential conflicts of interest (CoIs). This investigation: 1) evaluates the correspondence of two well-utilized CoI databases: the Centers for Medicare and Medicaid Services Open Payments (CMSOP) and ProPublica’s Dollars for Docs (PDD) and 2) quantifies CoIs among authors of a publically available point of care clinical support website. Two data sources were used: the hundred most common drugs and the top fifty causes of death. These topics were entered into a freely available database. The authors (N = 139) were then input into CMSOP and PDD and compensation and number of payment were determined for 2013-2015. The subset of highly compensated authors that also reported “Nothing to disclose” were further examined. There was a high degree of similarity between CMSOP and PDD for compensation (R^2^ ≥ 0.998) and payment number (R^2^ ≥ 0.992). The amount received was 1.4% higher in CMSOP ($4,059,194) than in PDD ($4,002,891). The articles where the authors had received the greatest compensation were in neurology (Parkinson’s Disease = $1,810,032), oncology (Acute Lymphoblastic Leukemia = $616,727), and endocrinology (Type I Diabetes = $377,388). Two authors reporting “Nothing to disclose” received appreciable and potentially relevant compensation. CMSOP and PDD produced almost identical results. CoIs were common among authors but self-reporting may be an inadequate reporting mechanism. Recommendations are offered for improving the CoI transparency of pharmaceutical-author interactions in point-of-care electronic resources.

---

Medscape is a medical reference site which can be used for medical advising and as a source of continuing medical education. The writers of these articles on diseases, symptoms, or drugs are health care professionals. Medscape was founded in 1995 as an online resource for better patient care for health professionals and interested consumers. It sought to provide peer-reviewed and edited information by renowned clinicians in fields such as AIDS, other infectious diseases, urology, and surgery (Frishauf, 2005). Medscape’s primary target was originally American clinicians but was available to anyone who registered for a free account. This open-door policy gave both physicians and laypersons equal access to healthcare related materials. Medscape currently provides medical news, journals, and Continuing Medical Education to healthcare professionals and information to non-healthcare professionals. If an author of a well-used, site is receiving funding from an outside company to advocate for a certain drug or treatment modality, it may put patients at risk for biased guidance or encouraging practices that are inconsistent with evidence based medicine. Medscape does provide some limited conflict of interest (CoI) disclosure information in terms of specific companies and involvement. However, this database does not disclose the quantity of remuneration. A $10 lunch may have a different potential for influence (Liu et al. 2017) than if compensation was sufficient to purchase a vacation, car, or new home.

The two databases that report pharmaceutical-physician interactions in the US are the Centers for Medicare and Medicaid Service’s Open Payments (CMSOP) and ProPublica’s Dollars for Docs (PDD). The Physician Payment Sunshine Act established the National Physician Transparency Program, also known as Open Payments as part of section 6002 of the Affordable Care Act and requires pharmaceutical companies and medical device manufacturers to report payments given to physicians or teaching hospitals (Marshall et al. 2016). It also discloses any ownership or investments held in US based medical companies. CMSOP was established in 2013 and includes a national database that shows the payments physicians have received from pharmaceutical and medical device companies. The accuracy of CMSOP was challenged in a study that determined that approximately one-quarter of doctors labeled as neurosurgeons were not (Babu et al. 2016). The PDD database was released by the nonprofit investigative journalism organization ProPublica in 2010 (Jones et al. 2016). PDD contains 9.2 billion in payments made to over nine-hundred thousand doctors by two-thousand companies (Tigas et al. 2018). The user interface differs slightly between PDD and CMSOP but PDD provides more CoI information including about specific products.

CoIs have previously been quantified for some influential materials (Liu et al. 2017). Three-quarters of the working groups responsible for writing the *Diagnostic and Statistical Manual of Mental Disorders* (DSM-5), the “bible” of psychiatry, were composed of members where the majority had ties to the pharmaceutical industry (Cosgrove & Krimsky, 2012). Although the dominant CoI framework is that the remuneration may precede potential influence, the reverse sequence may also occur. Advisors responsible for the approvals of the blood thinner ticagrelor (Brilanta) and the atypical antipsychotic quetiapine (Seroquel) received appreciable research support after being named to these government committees (Piller, 2018). Over one-quarter (26.2%) of *Goodman and Gilman’s Pharmacological Basis of Therapeutics*, a cornerstone pharmacology textbook in medical and pharmacy schools, had an undisclosed patent (Piper et al. 2016). Examination of six medical textbooks revealed 20 million unreported dollars according to PDD and 677 undisclosed patents (Piper et al. 2018). Inaccurate disclosures, defined as a failure to report receipt of > $5,000, were identified in 89.3% of clinical practice guidelines (Andreatos et al. 2017). A half-dozen conditions were searched in two subscription-based point-of-care databases and self-reported CoIs were common among authors and editors (Amber et al. 2014). However, no prior research has systematically examined publically available point of care computerized resources. Although it is assumed that CMSOP and PDD will produce equivalent results, the degree of correspondence between these databases is currently unknown.

Therefore, the objectives of this study were to: 1) determine whether PDD and CMSOP produce similar results, and 2) quantify the extent that Medscape authors received external funding from pharmaceutical and medical device companies. As some have found that self-reported CoI disclosure is inadequate (Andreatos et al. 2017), additional examination was made on disclosures of a subset of authors that reported “Nothing to disclose.”

## Methods

### Procedures

A list of the hundred most common drugs in the US was compiled (Supplementary Appendix) and an article pertaining to each was obtained in October, 2017. For example, for the #1 drug (levothyroxine), the corresponding entry was identified. Next, the top fifty causes of death compiled by the Centers for Disease Control and Prevention in the US in 2014 was entered into either Google or Medscape. If the search was conducted with Google, the cause was followed by “Medscape”. This technique assisted in screening for relevant Medscape articles. Next, the most descriptive and longest article was recorded along with the authors, co-authors, and editors. For example, one of the top causes of death is Prostate Cancer (Malignant), and this was matched to a Medscape article titled, “Metastatic and Advanced Prostate Cancer”. With the exception of #4-Unintentional Injury, #32-Disorder of Nonrheumatic Aortic Valve and #44 Kidney Failure (unspecified), this strategy was almost always (48/51 or 94.1%, note that there were two entries for “lung cancer-malignant”, non-small cell lung cancer and small cell lung cancer) successful in identifying a relevant entry. Each author’s or editor’s name was input into PDD and CMSOP. The three available full-years (2013, 2014, and 2015) were examined. The number of payments and amounts were recorded (and verified on March, 2018). The author’s gender was also recorded. The authors, co-authors, and editors were then searched on CMSOP. In the rare instance of multiple entries with the same first and last name, the listed location needed to be within commuting distance and the middle-initial and specialty (e.g. neurologist) were also examined. The years, the number of payments and payment amounts received were recorded. The medications and medical devices reported by PDD were further examined among authors or editors that reported “Nothing to disclose” as these discordant disclosures were of interest (Leo & Lacasse, 2009). All information reported is publically available. This study was approved by the Institutional Review Board of the Wright Center of Scranton.

### Data-analysis

A regression (R^2^) analysis was completed to determine the association between CMSOP and PDD. The null hypothesis was that there was no association between these databases. A paired t-test was completed to determine if the compensation was equivalent. GraphPrism was utilized for figure preparation. A *p* < .05 was considered statistically significant.

## Results

For the drug articles, the Medscape Drugs and Diseases Advisory Board (N = 8, 25.0% female) was analyzed with 75% having entries in PDD and CMSOP (Min = 1, Max = 31 payments; Min = $128, Max = $71,268). One author that wrote widely on oncology therapeutics reported “no relevant financial relationships” received $2,361 in 2014 with $1,195 for the chemotherapeutic agent everolimus (Afinitor) and $1,153 in 2015 with $995 for bevacizumab (Avastin). A chief editor that wrote entries on radiation related topics listed “nothing to disclose” but received $143,359 in 2015 and 2016, the preponderance (99.1%) from a company that produces radiation equipment (GE Healthcare). The 48 causes of death with relevant matching articles had 155 unique authors (18.1% female) with thirty authors contributing to multiple articles.

A regression analysis was completed to evaluate the correspondence between the CoI databases. The total number of payments reported in PDD and CMSOP were examined. The R^2^ values were 0.997, 0.998, and 0.992 in 2013, 2014, and 2015, respectively. The R^2^ values were 1.000, 0.999, and 0.998 in 2013, 2014, and 2015, respectively, for payment amounts (Supplemental Figure 1). These findings indicate a very high agreement between CMSOP and PDD for both measures. The total amount received was 0.016% or $590.23 higher in CMSOP ($3,684,068.23) than in PDD ($3,683,478) for 2013 to 2015. Figure 1 depicts the total amount received for authors for drugs and causes of death for each year and again shows substantial similarity between the CoI databases.

**Figure 1.**
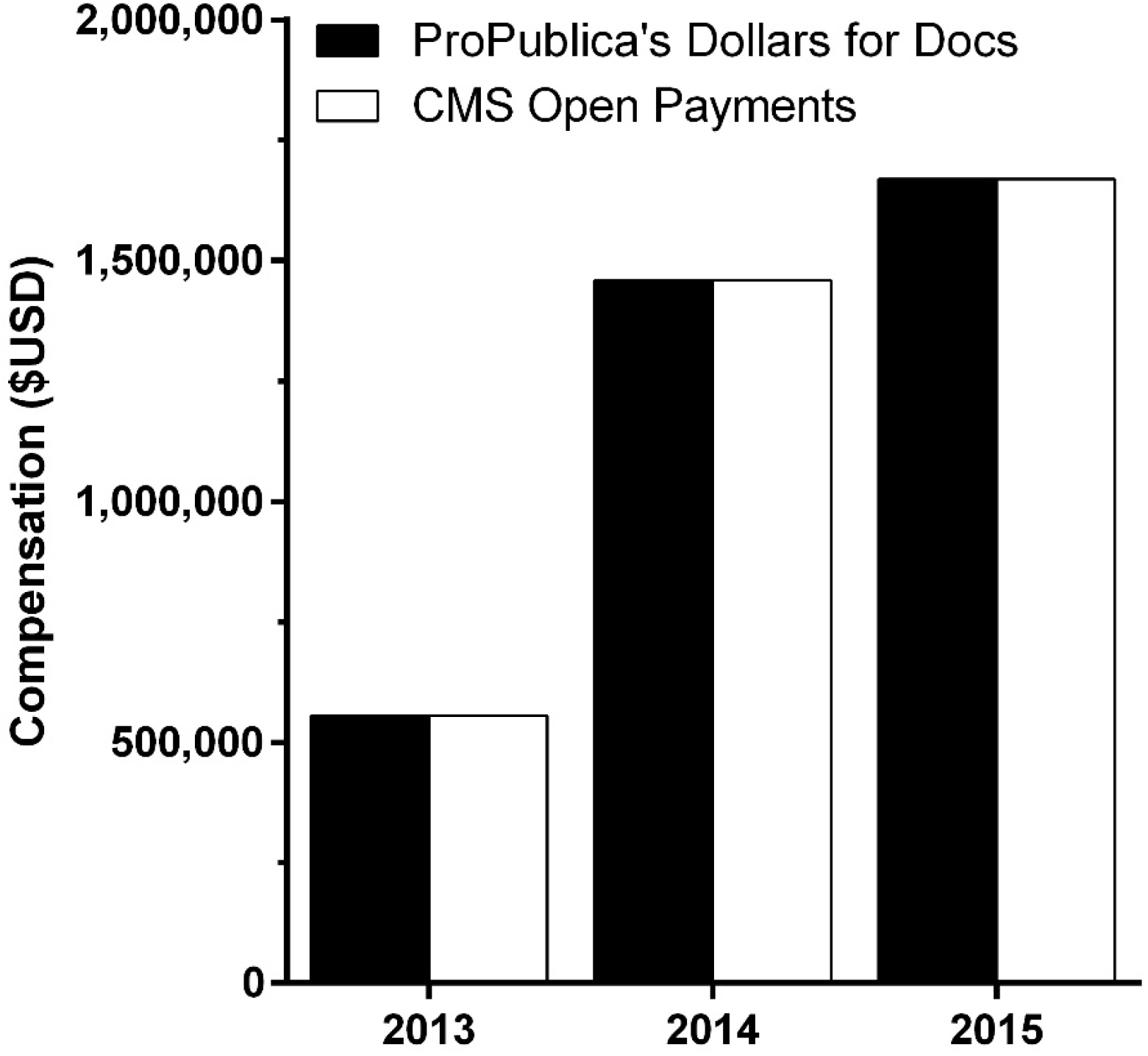
Total compensation for causes of death and drugs received by Medscape authors from 2013 to 2015 as reported by ProPublica’s Dollars for Docs and Center for Medicare and Medicaid Services (CMS) Open Payments conflict of interest databases.

Two-complementary analyses were completed. First, a ranking of the ten highest compensated articles, calculated as the sum of the compensation to one or more authors, is shown in Table 1. Oncology articles occupied six of the ten highest rankings. Females accounted for five of these 39 authors (12.8%). Additional examination was made with PDD among the authors who reported “Nothing to disclose”. A Parkinson Disease author received compensation for a device used in assessing Parkinson’s, Datscan Ioflupane ($4,071) and several Parkinson Disease pharmacotherapies (apomorphine: $15,462; carbidopa/levodopa: $17,721; and rasagiline: $29,521) in 2015. A Type I diabetes author, who again reported “Nothing to disclose”, received compensation for three drug treatments FDA approved for Type II diabetes: dapgliflozin ($75), liraglutide ($26,100), and exenatide ($128,000) in 2015.

**Table 1.** Top ten highest compensated articles of the top fifty US causes of death in Medscape as reported by the Centers for Medicare and Medicaid Services Open-Payments. Number of Open-Payments eligible authors or editors is shown in parentheses. *includes compensation from author reporting “Nothing to disclose”.

| Article | Compensation |
| --- | --- |
| 1. Parkinson Disease (3)* | \$1,810,032.07 |
| 2. Acute Lymphoblastic Leukemia (3) | \$616,726.90 |
| 3. Metastatic Cancer With Unknown Primary Site (5) | \$511,302.60 |
| 4. Multiple Myeloma (3) | \$489,735.01 |
| 5. Type 1 Diabetes Mellitus (2)* | \$377,657.24 |
| 6. Bladder Cancer (4) | \$336,763.58 |
| 7. Pancreatic Cancer (5) | \$269,896.99 |
| 8. Breast Cancer (2) | \$256,677.01 |
| 9. Dilated Cardiomyopathy (8) | \$187,291.78 |
| 10. Chronic Obstructive Pulmonary Disease (5) | \$174,669.87 |

Second, disclosures were further examined among authors responsible for entries on the top ten causes of death (N =31). Three-fifths of these authors (61.3%) were discordant in that they self-reported no disclosures but had a PDD entry (total = $459,967 although 95.5% was from two-authors including the diabetes author mentioned previously). Two-thirds of discordant authors (68.4%) received less than $1,000 and the vast majority (84.2%) received less than $5,000. A non-small cell lung cancer author that reported no relevant CoIs to Medscape was the recipient of $62,536 including $8,839 for the chemotherapy palbociclib (Abraxane), $4,285 for pembrolizumab (Keytruda), and $2,365 for crizotinib (Xalkori) in 2015.

Even among all authors with disclosures, there were appreciably less information provided in their disclosures than in PDD. For example, a neurologist who co-authored the Parkinson Disease entry listed only consulting when PDD noted $45,800 in Honoraria and $13,808 in travel and lodging in 2014 alone. PDD included $35,680 in 2015 for rasagiline but the corresponding manufacturer was not included in the Medscape disclosure.

## Discussion

There are several key findings in the study. Most authors reporting remuneration received from pharmaceutical companies or medical device manufacturers did so accurately. However, inconsistencies were occasionally noted. This discrepancy could result from the authors correctly interpreting that their compensation was not relevant to the article. However, self-reporting CoI may in some cases also be inadequate (Andreatos et al. 2017). Thirty-percent of the National Academy of Sciences report on genetically engineered crops omitted a COI including research funding and patents (Krimsky & Schwab, 2017). Alternatively, the compensation may have been received outside of the temporal window of reporting. The time-frame is challenging to determine because articles only provide info on when an article was updated but not when it was originally written.

PDD provides a substantial level of detail about the specific medications where authors received compensation. The neurologist who wrote the Parkinson’s article received over one-hundred thousand dollars from companies related to Parkinson’s treatments and diagnosis. However, the majority of this remuneration was related to antiepileptic drugs. The endocrinologist who wrote the diabetes article received the preponderance of this compensation for anti-diabetes pharmacotherapies. It is currently ambiguous whether compensation for a medicine for Type II diabetes would, or even should, be reported in an article on Type I diabetes.

This investigation showed that PDD and CMSOP produced equivalent results in the number and amount payments. Similarly, both PDD and CMPOP produced near identical outcomes in determining whether an entry was present. This consistency may at least partially offset earlier criticisms (Babu et al. 2016). These databases just differ slightly in their interface and timeliness of updating. One minor difference is that PDD rounds the payment to the dollar, whereas CMSOP shows the cent values as well. Another difference is that CMSOP provides more recent information than PDD. Because PDD and CMSOP are congruent with each other, this shows both databases have reliably compiled pharmaceutical-provider transactions. Future quantitative bioethics research with a larger and more diverse (e.g. more general practitioners) sample of physicians is needed to determine if PDD and CMSOP are functionally equivalent.

Although not the main focus of this study, it did not escape notice that the vast majority of authors of this influential content were male. The preponderance (88.9%) of the authors of the ten most highly remunerated articles were male. Similarly, eight out of every nine authors of a widely used pharmacology textbook, *Goodman and Gilman’s Pharmacological Basis of Therapeutics*, were male (Piper et al. 2016). A substantial under-representation of female authors were also observed for pathophysiology textbooks used to train allopathic or osteopathic physicians (Piper et al. 2018). Males accounted for two-thirds of the recipients of the 2.4 billion in industry payments to US physicians. The payment per physician was over-three fold larger for males ($5,031) then females ($1,390) (Triagle et al. 2017). Collectively, these findings point toward pronounced sex differences in physician-industry interactions.

There is an awareness that the identification of inaccurately reported CoIs can be a sensitive. The Leo-DeAngelis Lexapro episode involved a journal editor resisting transparency and engaging in behaviors more appropriate of a public relations firm than of a leader in evidence based medicine (DeAngelis & Fontanarosa, 2009; Leo & Lacasse, 2009; Melnick & Fugh-Berman, 2009). The intent of this report was not to impugn the reputation of any individual or organization but instead to be able to explore CoI disclosure accuracy and more critically evaluate the presented information. If there were multiple authors who self-report “nothing to disclose” who are found to have received appreciable, recent and relevant compensation, perhaps the point of care database should consider a direct link to the PDD report. Failures to report CoI (e.g. Jose Baselga of Memorial Sloan Kettering Cancer) raises appreciable concerns about the integrity of preclinical and clinical trial information (Miller, 2018). Authors and editors at point of care information resources that engage in this practice risk impugning the faith of the public and medical community in the integrity of their employers and should be subject to corrective measures.

Three key limitations should be noted. Since only physicians are searchable in CoI databases, pharmacists, scientists and nurse authors were not recorded nor compared. There is currently not a thorough understanding of industry interactions among non-physicians. This may change beginning in 2022 with the national inclusion of physician assistants and advanced practice nurses (Stewart, 2018). Second, the presence of potential CoI were appreciable. However, the goal of this study was not to determine whether CoIs impacted information presentation. This medical information database is presumed to be an unbiased site that the general public and clinicians use for medical advising. It reaches a wide audience in part due to its nature as a freely available internet resource. If an author of a well-used site is receiving funding from an outside company to advocate for a certain drug or treatment, this may put clinicians and patients at risk of misguidance or receiving information that is incongruent with evidence-based medicine. Self-reported CoIs, or their absence, should be interpreted with caution. There is some subjectivity in what information to report. For example, the author of an article on Type I diabetes may not believe that compensation related to a drug for Type II diabetes is relevant. The International Committee of Medical Journal Editors (2010) indicates that “You should disclose interactions with ANY entity that could be considered broadly relevant to the work”. Third, the time-frame covered for disclosures is also important. Because the database only provided information on when an article was updated, but not originally written, we can not exclude the possibility that the original authorship may have preceded the reported remuneration. However, as there are many biomedical developments, we can only hope that the preponderance of content was from post-2012. Further, influential parties have been known to be compensated in what appears to be a services first/pay-later pattern (Piller, 2018).

In conclusion, given the prevalence of potential CoIs, and occasional instances of inaccuracies, further research systematically evaluating other point of care online resources (e.g. UpToDate) may be warranted. At least based on this one database and examination of the ten highest remunerated articles, self-reported financial CoIs were generally verified with PDD. This finding contradicts some prior investigations (21, 28). However, the inconsistencies, while uncommon, suggest there is at least some potential for improved transparency. Suggestions for point of care databases would be to develop shared standards regarding the time period of reporting, a precise definition of “relevant” activities which would result in disclosures, reporting at least the categories of compensation (e.g. $10-9,999; $10,000-99,999; ≥ $100,000), and developing procedures to mitigate potential bias. Modifications like these will insure that public and health care provider trust in the integrity of these resources is preserved.

## Data Availability

The data is available as a supplemental file.

## Acknowledgements

Software to complete this research was provided by the National Institute of Environment Health Sciences (NIEHS T32-ES007060-31A1). No specific funding was received for this study. We thank ProPublica for making the Dollars for Docs database publically available. An earlier version of this paper was completed as part of course requirement for Readings in Biomedical Sciences with Prof. Darina Lazarova.

---

**Supplemental Figure 1.**
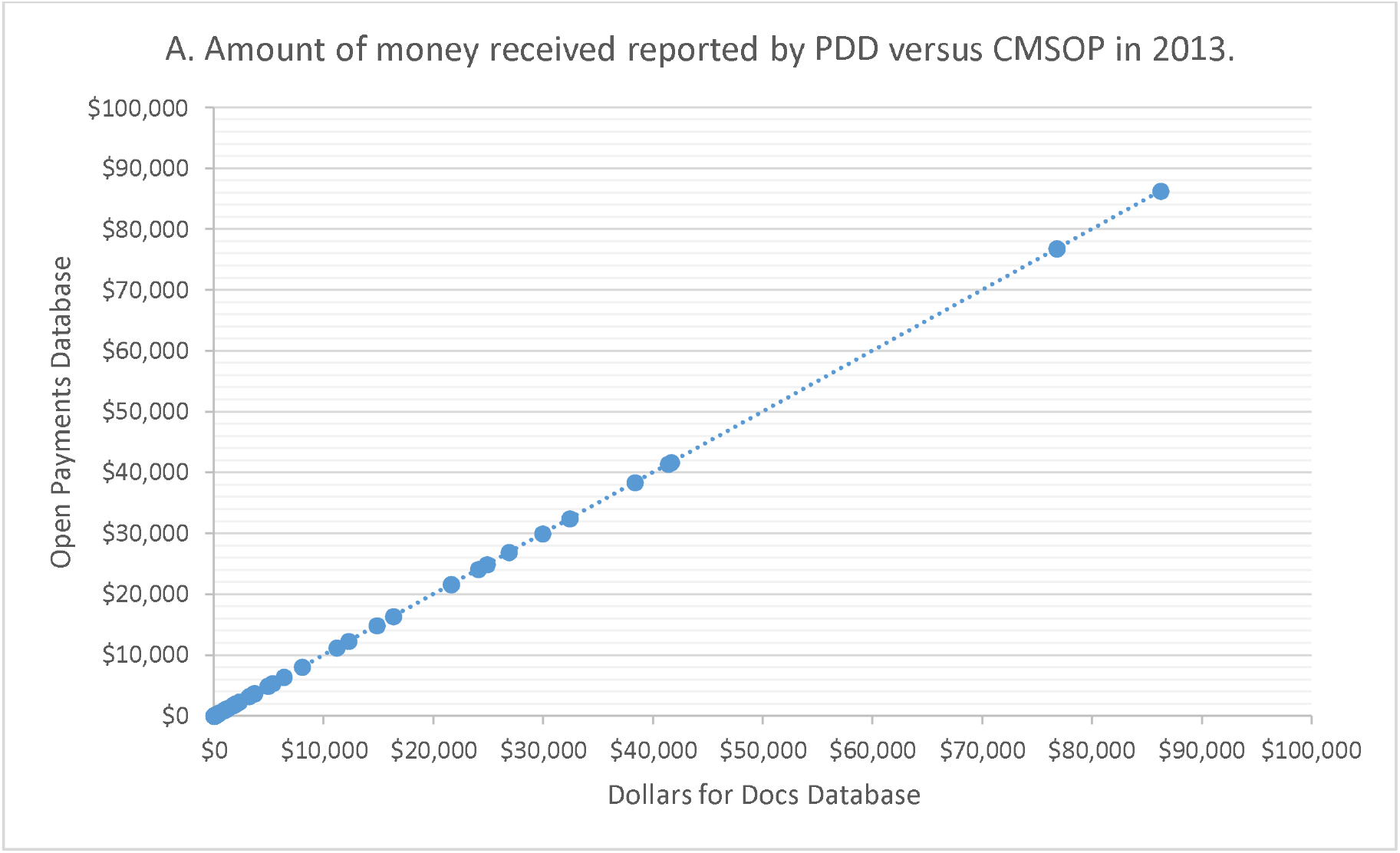
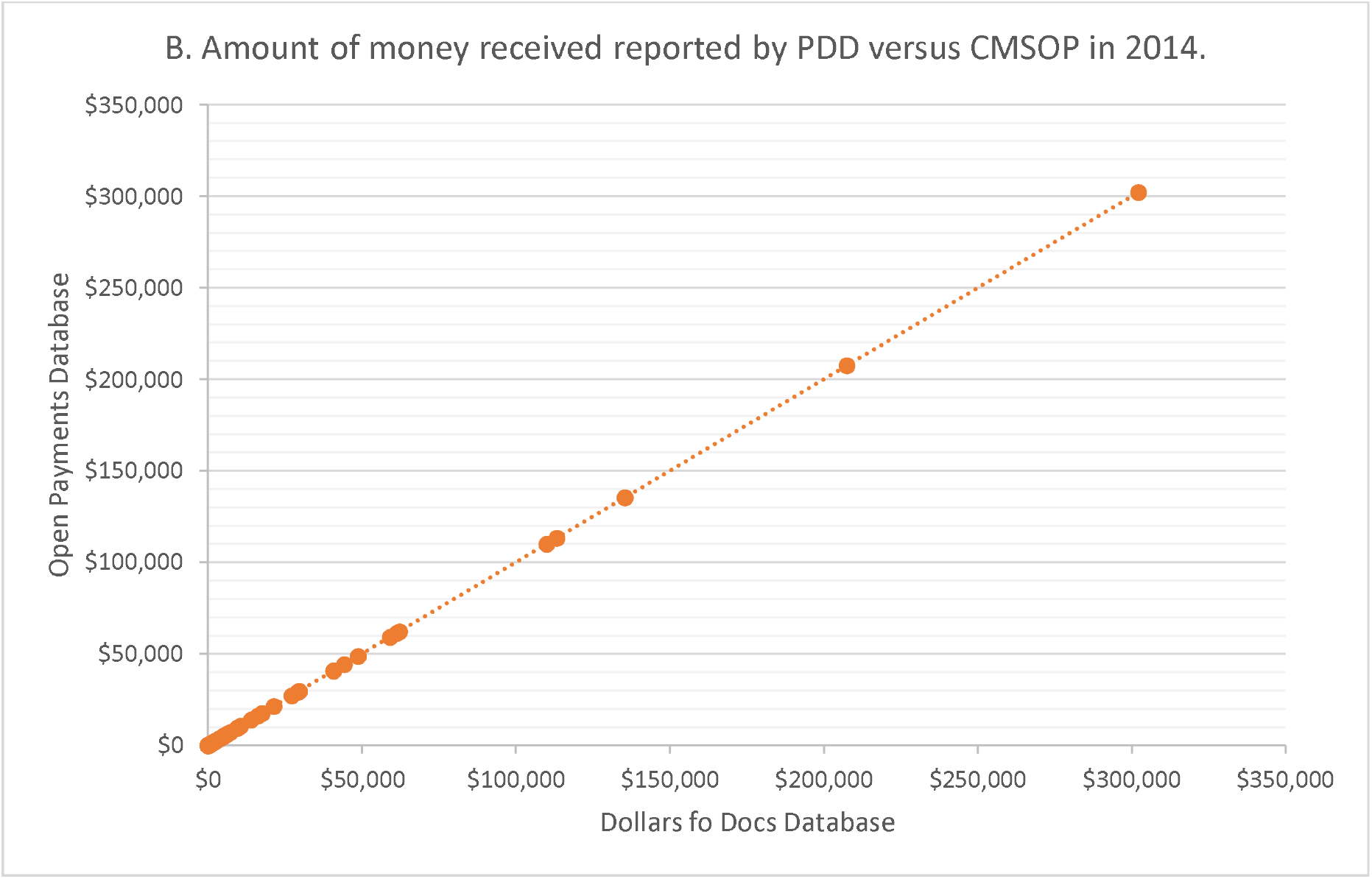
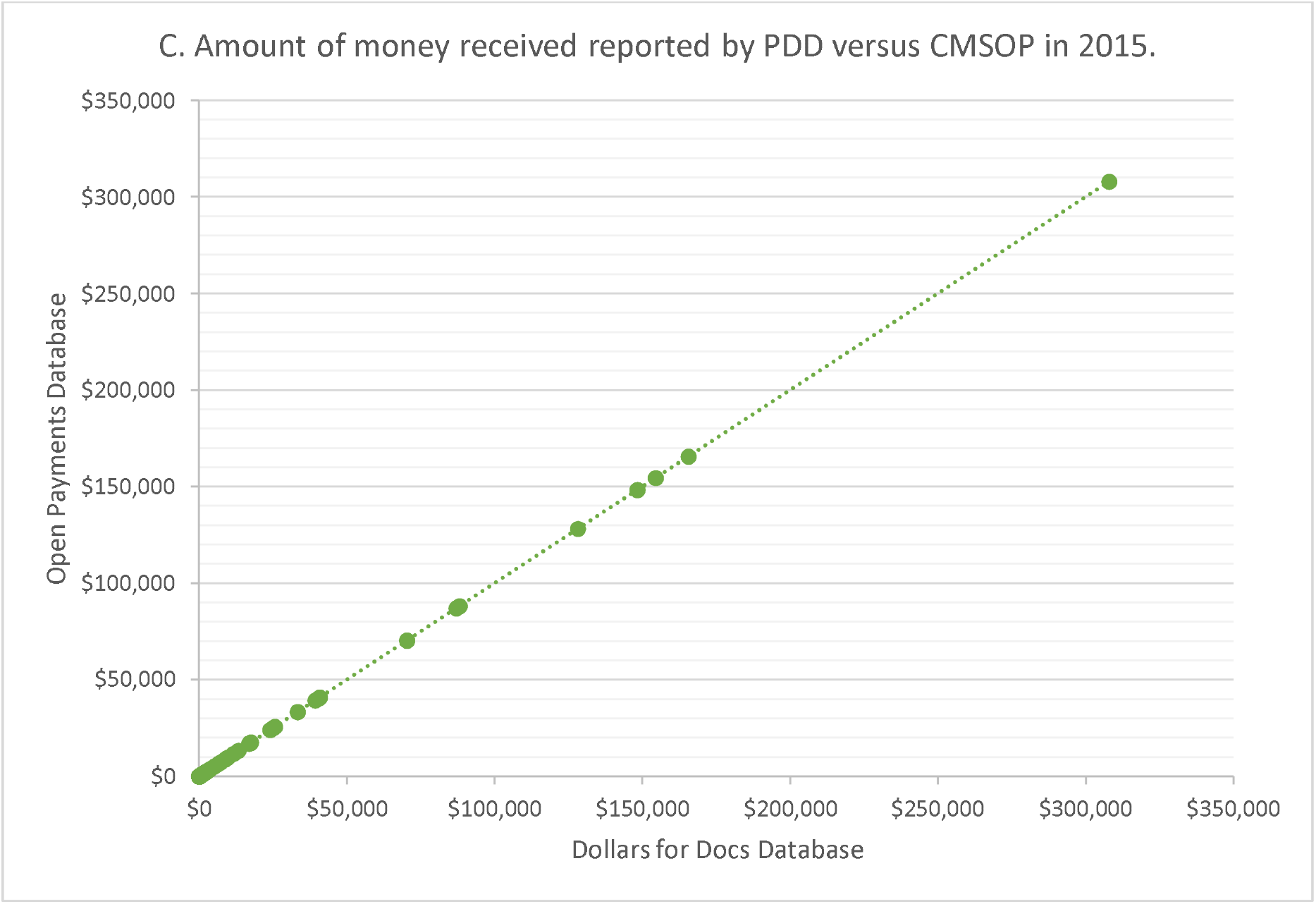
The total payments received by Medscape writers from years 2013 (A), 2014 (B) and 2015 (C) as reported by ProPublica’s Dollars for Docs (PDD) and Center for Medicare and Medicaid Services Open Payments (CMSOP).

